# First-Day Documentation Patterns and Mortality or Continued ICU Occupancy Across Four Critical Care Datasets: A Fixed-Window Retrospective Study

**DOI:** 10.64898/2026.06.04.26354922

**Authors:** Alexis M. Collier

## Abstract

**Background:** Documentation patterns may contain contextual information about patient complexity, monitoring, workflow, and local electronic health record practice. Different analytic windows support different research purposes. Broad encounter-level summaries can characterize documentation and workflow across a stay, while a fixed early window is needed to study whether information available at a defined landmark is associated with later outcomes.

**Objective:** To evaluate whether documentation volume, diversity, timing, and availability during the first 24 ICU hours provide retrospective information about post-landmark outcomes across four critical care datasets.

**Methods:** We conducted separate fixed-window analyses in NWICU, eICU, a nursing-chart and laboratory-density dataset, and HiRID. Predictors were restricted to information available by hour 24. Models used grouped or stay-level out-of-fold validation. Results were not pooled because cohorts, endpoints, feature semantics, and comparators differed.

**Results:** In NWICU, 21,609 eligible stays were evaluated for ICU occupancy beyond hour 72. A fixed-window enhanced model achieved AUROC 0.689 versus 0.527 for a care-unit baseline, a paired difference of 0.162 (95% CI 0.155 to 0.176). In eICU, 5,107 deduplicated stays with 465 hospital deaths were evaluated. Age and sex plus first-day documentation features achieved AUROC 0.6712 versus 0.6073 for age and sex, a difference of 0.0639 (95% CI 0.0382 to 0.0905). In the nursing-chart dataset, first-day features improved observed follow-up mortality AUROC from 0.659 to 0.721 and post-landmark long-stay AUROC from 0.546 to 0.726. In HiRID, the strict first-day combined model achieved AUROC 0.645; documentation-only AUROC was 0.644 and demographics-only AUROC was 0.532. Label-permutation controls were near random where available.

**Conclusions:** First-day documentation patterns showed heterogeneous, retrospective association with later mortality or continued ICU occupancy. Effect size depended on the endpoint, comparator, timestamp semantics, and feature family. The findings support documentation as a contextual input for further fixed-window and silent-mode research. They do not establish causal workload measurement, staffing need, clinical benefit, transportability, or deployment readiness.

## 1. Introduction

Electronic health records capture values and the processes through which care is documented. Counts of charted events, source diversity, timing concentration, gaps, and stream presence can reflect patient complexity, monitoring intensity, clinician response, local policy, interface design, and extraction rules [1-8]. These features are attractive for clinical artificial intelligence because they can be derived from routine data without adding documentation tasks. Their interpretation is difficult because the same patterns may reflect patient state, care delivery, workflow, or data-system behavior.

Temporal design follows the intended question. Encounter-level documentation summaries are appropriate for descriptive and retrospective workflow characterization. Prospective-style prognostic questions require a declared landmark, information available by that time, patient-level or stay-level separation, and outcome follow-up beginning after the landmark [9-11]. Version 2 adopts the latter design so the four datasets can be examined under one shared first-day framework.

Version 1 examined a broader portfolio of documentation-density, workflow, and long-stay proxy analyses with dataset-specific windows. The present study intentionally narrows the scope to independently reconstructed first-24-hour analyses in four datasets. Its objective is to determine whether early documentation patterns show reproducible retrospective signal and to define the boundaries of that signal across mortality and continued-occupancy outcomes. Because the purpose and estimand changed, Version 2 establishes new cohorts and does not treat its estimates as one-to-one replacements for the earlier analyses.

## 2. Methods

### 2.1 Study design

This was a retrospective, multi-dataset fixed-window evaluation. Each analysis used a 24-hour landmark and retained only predictors available by that landmark. Dataset-specific models, endpoints, and uncertainty procedures were preserved. Estimates were not pooled because the analyses differed in cohort construction, outcome definition, feature semantics, missingness, and comparator strength.

### 2.2 Datasets and outcomes

NWICU used ICU-stay and chart-event extracts [1]. Eligible stays remained in the ICU beyond hour 24. The outcome was continued ICU occupancy beyond hour 72, equivalent to more than 48 additional hours after the landmark. Mortality was not analyzed because the local source package lacked a defensible mortality field.

eICU used a prepared modeling cohort and raw nurseCharting, intakeOutput, and infusionDrug streams [2,3]. The outcome was hospital mortality among stays represented through hour 24. Exact duplicate cohort rows were removed before modeling.

The nursing-chart analysis used dtNursingChart and dtLab extracts with empirically interpreted admission-relative offsets. Outcomes were observed follow-up mortality, for which the exact horizon was unavailable, and ICU occupancy beyond hour 168 among patients still in the ICU at hour 24.

HiRID used a derived table containing explicitly named first-24-hour observation counts [4,5]. The primary outcome was ICU mortality among stays lasting at least 24 hours. Raw HiRID event shards were unavailable, so this analysis is a derived-table reanalysis rather than a raw-source reconstruction.

**Table 1.** Fixed-window cohorts and outcomes.

| Dataset | Analysis N | Outcome events | Outcome | Primary comparator |
| --- | --- | --- | --- | --- |
| NWICU | 21,609 stays | 9,068 | ICU occupancy beyond hour 72 | Care unit |
| eICU | 5,107 stays | 465 | Hospital mortality | Age and sex |
| dt nursing-chart mortality | 1,873 patients | 1,016 | Observed follow-up mortality | Available baseline variables |
| dt nursing-chart long stay | 2,356 patients | 965 | ICU occupancy beyond hour 168 | Available baseline variables |
| HiRID | 15,384 stays | 1,297 | ICU mortality | Age and sex |

### 2.3 Feature construction

NWICU features were reconstructed from events charted between ICU entry and hour 24 and available in storage by the landmark. Full-stay counts, full-stay spans, last full-stay chart offset, total length of stay, outtime, and post-landmark events were prohibited. The enhanced feature set combined first-day chart-event and vital-sign patterns; it should therefore be interpreted as a mixed workflow and physiology model rather than documentation alone.

eICU features were rebuilt from offsets 0 through 1,440 minutes. They included event counts, unique labels or drugs, four six-hour bins, stream-observed indicators, active documentation time, gaps, gap variation, burstiness, and boundary timing. Physiologic values, discharge fields, ICU length of stay, mortality labels, and post-window events were excluded.

The nursing-chart models used first-day nursing and laboratory features with no negative or post-landmark events. Forbidden discharge and outcome features were excluded. HiRID used admission age and sex, log-transformed number of observations during the first 24 hours, and number of variables represented during the first 24 hours. A mapped feature labeled idi_events_24h failed temporal audit and was excluded because it behaved like a full-stay count.

### 2.4 Modeling and validation

Analyses used logistic models with preprocessing performed inside training folds. NWICU used repeated grouped cross-validation with patient-level separation. eICU used five repeats of five-fold stratified stay-level cross-validation. HiRID used ten repeats of five-fold stratified out-of-fold prediction. The nursing-chart analysis used patient-level out-of-fold validation. AUROC was the primary discrimination measure. Brier score, log loss, calibration intercept and slope, bootstrap intervals, and label-permutation controls were reported where available. The eICU and HiRID bootstrap procedures quantified sampling uncertainty conditional on saved out-of-fold predictions rather than refitting every model in each replicate.

## 3. Results

### 3.1 NWICU

The NWICU analysis included 21,609 eligible stays from 18,089 patients. There were 9,068 continued-occupancy events (41.96%). A care-unit baseline achieved AUROC 0.527 (95% CI 0.518 to 0.534), Brier score 0.2428, and calibration slope 0.894. The fixed-window enhanced model achieved AUROC 0.689 (95% CI 0.685 to 0.699), Brier score 0.2164, and calibration slope 1.135. The paired AUROC increase was 0.162 (95% CI 0.155 to 0.176). Results were stable across repeated grouped validation. Label-shuffle AUROCs were 0.494 for the baseline and 0.504 for the enhanced model.

The timestamp audit retained 3,205,286 chart events that were both clinically recorded and stored by hour 24. It excluded 86,215 observations charted before the landmark but stored afterward. No post-landmark events were used. A total of 6,753 eligible stays (31.25%) had no available chart event by the landmark. The model therefore demonstrates internal association with future ICU occupancy, not mortality, workload, or transportability.

### 3.2 eICU

The 9,397-row source cohort represented 5,107 unique ICU stays; 4,290 rows were exact duplicates and were removed. The final cohort contained 465 hospital deaths (9.11%). The age-and-sex baseline achieved AUROC 0.6073 (95% CI 0.5791 to 0.6351) and Brier score 0.08184. Adding all first-day documentation features increased AUROC to 0.6712 (95% CI 0.6432 to 0.6970) and improved Brier score to 0.07992. The paired AUROC increase was 0.0639 (95% CI 0.0382 to 0.0905). Calibration intercept and slope were -0.333 and 0.847 for the enhanced model.

Feature-family analysis showed complementarity rather than strong standalone prediction.

Documentation-only AUROC was 0.6352. A volume, diversity, and activity family achieved combined AUROC 0.6740 versus 0.6073 for age and sex, a difference of 0.0668 (95% CI 0.0426 to 0.0908). Timing and concentration features alone achieved AUROC 0.5943, while the combined model achieved 0.6423. Nurse-rhythm features alone achieved AUROC 0.5896, while the combined model achieved 0.6401. Across 100 mortality permutations, mean enhanced AUROC was 0.4975.

The raw-event audit excluded 3,511,317 post-window nurseCharting rows, 293,543 post-window intakeOutput rows, and 141,761 post-window infusionDrug rows. Intake/output and infusion streams were absent during the first day for 580 and 3,156 stays. Missingness indicators alone achieved AUROC 0.5296 and added no meaningful increment to quantitative documentation features.

### 3.3 Nursing-chart and laboratory-density dataset

The mortality cohort included 1,873 patients with explicit Alive or Death follow-up status, including 1,016 deaths (54.2%). The baseline achieved AUROC 0.659 (95% CI 0.634 to 0.683), and the enhanced first-day model achieved 0.721 (95% CI 0.696 to 0.743). The paired difference was 0.062 (95% CI 0.043 to 0.082). Brier score improved from 0.229 to 0.211; enhanced calibration slope was 0.87. Because the mortality horizon was unavailable, this result must be described as observed follow-up mortality rather than in-hospital or fixed-horizon mortality.

The long-stay cohort included 2,356 patients still in the ICU at hour 24, of whom 965 remained beyond hour 168. Baseline AUROC was 0.546 (95% CI 0.522 to 0.570), and enhanced AUROC was 0.726 (95% CI 0.706 to 0.745), a difference of 0.180 (95% CI 0.152 to 0.206). Brier score improved from 0.241 to 0.206. Terminal ICU disposition was unavailable, creating a competing-event limitation. The exact timestamp origin was empirically supported but not confirmed by a source data dictionary.

### 3.4 HiRID

The primary HiRID cohort included 15,384 stays lasting at least 24 hours, with 1,297 ICU deaths (8.43%).

The combined age, sex, and two explicitly first-day documentation-count model achieved AUROC 0.645 (95% CI 0.632 to 0.658), AUPRC 0.125 (95% CI 0.118 to 0.136), Brier score 0.0757, calibration intercept - 0.040, and calibration slope 0.983. Documentation-only AUROC was 0.644, while demographics-only AUROC was 0.532. Thus, most discrimination came from early documentation volume and breadth rather than age and sex.

The all-stay sensitivity achieved AUROC 0.629, and the at-least-48-hour sensitivity achieved 0.610, indicating cohort dependence. Across 100 label-shuffle refits, mean AUROC was 0.498 and the maximum was 0.532. The mapped idi_events_24h feature was excluded because it disagreed with the explicitly first-day count in every stay lasting at least 24 hours, with a median excess of 3,131 events. Raw source shards were unavailable, and repeat admissions from the same person could not be ruled out.

**Table 2.** Primary fixed-window performance results.

| Dataset and endpoint | Baseline AUROC | Enhanced AUROC | Difference, 95% CI | Key boundary |
| --- | --- | --- | --- | --- |
| NWICU, occupancy beyond h72 | 0.527 | 0.689 | +0.162, 0.155 to 0.176 | Mixed chart and vital-sign model |
| eICU, hospital mortality | 0.6073 | 0.6712 | +0.0639, 0.0382 to 0.0905 | Cohort represented through h24 |
| dt, observed follow-up mortality | 0.659 | 0.721 | +0.062, 0.043 to 0.082 | Mortality horizon unavailable |
| dt, occupancy beyond h168 | 0.546 | 0.726 | +0.180, 0.152 to 0.206 | Competing-event status unavailable |
| HiRID, ICU mortality | 0.532 | 0.645 | Not estimated as paired primary contrast | Derived-table reanalysis |

## 4. Discussion

### 4.1 Principal findings

Across four critical care datasets, first-day documentation patterns showed retrospective association with later mortality or continued ICU occupancy. The direction was generally positive, but the meaning and magnitude varied substantially. NWICU and the nursing-chart long-stay analysis showed larger increments for post-landmark occupancy outcomes. eICU showed a modest mortality increment over age and sex. HiRID showed modest discrimination in which first-day documentation counts carried nearly all of the signal. These estimates should not be compared as a leaderboard because endpoints, baselines, feature families, and data provenance differed.

The fixed-window analyses support a focused interpretation that complements the broader purpose of Version 1. Documentation patterns can act as contextual inputs in an early-window prognostic design, but they are not direct measures of workload, staffing adequacy, documentation quality, clinician vigilance, or patient instability. Event volume may reflect monitoring, acuity, policy, treatment, interface design, and extraction. Timing patterns may reflect workflow or storage delay. Missing streams may reflect absent care, absent documentation, or absent extraction. Construct interpretation requires independent ground truth.

### 4.2 Relationship between analytic purpose and temporal design

The appropriate feature window depends on the research purpose. Version 1 used broader documentation-density and workflow analyses to study encounter-level and long-stay proxy patterns across heterogeneous datasets. Version 2 asks a distinct prognostic question under a shared first-day landmark. Information available by hour 24 is therefore paired with outcomes beginning after that landmark. This design aligns cohort eligibility, predictors, validation, and interpretation with the newly defined estimand.

Implementing the common first-day design required dataset-specific reconstruction. NWICU distinguished charttime from storetime to determine availability at the landmark. eICU established one analytic row per unique stay and reconstructed features from raw event offsets. HiRID used fields explicitly defined for the first 24 hours. These decisions were necessary for the focused Version 2 question and complement the broader descriptive and proxy-oriented purpose of Version 1.

### 4.3 Calibration, comparison, and transport

Discrimination alone is insufficient. The eICU enhanced calibration slope of 0.847 indicated over-extreme internal risks despite mean predicted mortality matching prevalence. The HiRID combined model calibrated well internally but was built from a derived table and was not externally transferred. The nursing-chart mortality slope of 0.87 also warrants external calibration assessment. Each dataset requires a clinically appropriate same-time comparator. Age and sex are limited; future mortality studies should test whether documentation adds value beyond physiology, severity, comorbidity, and observable clinician response.

Cross-site transport cannot be inferred from four separate within-dataset analyses. Documentation semantics depend on source tables, timestamp conventions, unit policy, staffing, and EHR configuration. A common feature label does not guarantee semantic parity. External validation requires locked definitions, identical availability rules, aligned endpoints, and transparent missingness handling.

### 4.4 Implications for nursing informatics and clinical AI

The findings justify prospective silent-mode research, not active deployment. A next study should declare the intended user, decision, prediction time, outcome horizon, action pathway, and prohibited uses. It should combine documentation features with a strong clinical comparator, preserve patient-level separation, assess calibration and subgroup uncertainty, and evaluate decision-relevant utility at a prespecified alert budget.

Workflow outcomes should include nurse-reported burden, observed task timing, staffing context, documentation delay, and independent reliability adjudication. Documentation features should not be used for individual productivity ranking or punitive workforce decisions without validated constructs and governance safeguards [6-8,11-13].

### 4.5 Limitations

- All analyses were retrospective and internally validated. No model was evaluated in prospective silent mode or active clinical use.
- Cohorts, endpoints, feature definitions, and baselines differed, preventing pooled inference or direct performance comparison.
- The NWICU enhanced model combined documentation and vital-sign features and therefore does not isolate documentation-specific value.
- The eICU cohort was conditioned on representation through 24 hours and could not assess early deaths or discharges.
- The nursing-chart mortality horizon and exact timestamp dictionary were unavailable; the long-stay analysis lacked terminal-disposition information.
- The HiRID analysis used a derived table without raw event shards or a separate longitudinal subject identifier.
- Subgroup, treatment, staffing, benefit, harm, and decision-utility outcomes were incomplete or unavailable.
- Bootstrap intervals in some analyses were conditional on fixed out-of-fold predictions rather than complete model refitting.

## 5. Conclusion

Fixed first-day documentation patterns carried heterogeneous retrospective signal across four critical care datasets. Temporally valid models produced moderate rather than extraordinary performance. The strongest defensible interpretation is that documentation volume, diversity, timing, and availability may complement clinical context for later mortality or continued-occupancy modeling. The evidence does not establish causal workload measurement, staffing need, clinical benefit, external transportability, or deployment readiness.

Future work should prioritize locked temporal contracts, strong same-time comparators, raw-source reproducibility, external validation, subgroup calibration, and prospective nonpunitive workflow evaluation.

## Data Availability

This manuscript reports aggregate retrospective analyses derived from de-identified and controlled-access critical care datasets. Public datasets discussed in this manuscript are available through PhysioNet or their respective repositories subject to credentialing, required training, data-use agreements, and dataset-specific access restrictions. The author cannot redistribute patient-level data, restricted tables, raw extracts, or derived row-level files from controlled-access datasets. Aggregate results needed to interpret the manuscript are included in the text and tables. Code is available from the author upon reasonable request, subject to removal of local paths, credentials, protected dataset structure details, and any material restricted by data-use agreements.

https://physionet.org/content/nwicu-northwestern-icu/0.1.0/

https://physionet.org/content/mimiciv/

https://physionet.org/content/mimic-iv-note/

https://physionet.org/content/eicu-crd/2.0/

https://physionet.org/content/hirid/1.1.1/

## Ethics Statement

This study used de-identified retrospective datasets and aggregate validation outputs. Public datasets were accessed under their respective credentialing, training, and data-use requirements. No prospective intervention was conducted, no patients were contacted, and no identifiable patient information is reported.

## Data Availability

NWICU, eICU, MIMIC-derived resources, and HiRID are available through PhysioNet subject to credentialing, training, data-use agreements, and dataset-specific restrictions [1-5]. Patient-level and restricted derived files are not redistributed. Aggregate results needed to interpret this manuscript are reported in the text and tables.

## Code Availability

Reproducible analysis scripts, provenance manifests, aggregate outputs, and verification artifacts are preserved by the author. A public code package may be released after removal of local paths and review for compliance with controlled-access dataset terms.

## Author Contributions

Alexis Collier conceived the study, developed or supervised the analyses, audited the evidence, interpreted the findings, drafted and revised the manuscript, and approved the final version.

## Related Presentation and Preprint Overlap

The fixed-window eICU analysis reported here is also included in a revised paper submitted to the Pacific Symposium on Biocomputing 2027. That conference paper presents the eICU component as a focused study. It is not an independent replication of the eICU analysis in this preprint. This manuscript presents the broader four-dataset fixed-window evaluation.

